# COVID-19 and homelessness in England: a modelling study of the COVID-19 pandemic among people experiencing homelessness, and the impact of a residential intervention to isolate vulnerable people and care for people with symptoms

**DOI:** 10.1101/2020.05.04.20079301

**Authors:** Dan Lewer, Isobel Braithwaite, Miriam Bullock, Max T Eyre, Robert W Aldridge, Alistair Story, Andrew Hayward

## Abstract

**Background:** There is an ongoing pandemic of the viral respiratory disease COVID-19. People experiencing homelessness are vulnerable to infection and severe disease. Health and housing authorities in England have developed a residential intervention that aims to isolate those vulnerable to severe disease (COVID-PROTECT) and care for people with symptoms (COVID-CARE).

**Methods:** We used a discrete-time Markov chain model to forecast COVID-19 infections among people experiencing homelessness, given strong containment measures in the general population and some transmission among 35,817 people living in 1,065 hostels, and 11,748 people sleeping rough (the ’do nothing’ scenario). We then estimated demand for beds if those eligible are offered COVID-PROTECT and COVID-CARE. We estimated the reduction in the number of COVID-19 cases, deaths, and hospital admissions that could be achieved by these interventions. We also conducted sensitivity and scenario analyses to identify programme success factors.

**Results:** In a ’do nothing’ scenario, we estimate that 34% of the homeless population could get COVID-19 between March and August 2020, with 364 deaths, 4,074 hospital admissions and 572 critical care admissions. In our ’base intervention’ scenario, demand for COVID-PROTECT peaks at 9,934 beds, and demand for COVID-CARE peaks at 1,366 beds. The intervention could reduce transmission by removing symptomatic individuals from the community, and preventing vulnerable individuals from being infected. This could lead to a reduction of 164 deaths, 2,624 hospital admissions, and 248 critical care admissions over this period. Sensitivity analyses showed that the number of deaths is sensitive to transmission of COVID-19 in COVID-PROTECT. If COVID-PROTECT capacity is limited, scenario analyses show the benefit of prioritising people who are vulnerable to severe disease.

**Conclusion:** Supportive accommodation can mitigate the impact of the COVID-19 pandemic on the homeless population of England, and reduce the burden on acute hospitals.

## Introduction

The pandemic of the respiratory disease COVID-19 is a threat to the health of populations around the world.^1^ People experiencing homelessness are particularly vulnerable due to the potential for outbreaks in multi-occupancy temporary accommodation, barriers to hand and respiratory hygiene, and comorbidities that increase the likelihood of severe COVID-19 disease. The size and structure of the homeless population is difficult to estimate, not least because there are different types of homelessness. One estimate suggests there are 143,000 people in ’core homelessness’ groups in England, which includes people sleeping rough, living in tents or cars, squatting, living in temporary homeless hostels, and sofa-surfing.^2^

COVID-19 severity is associated with pre-existing health conditions including cardiovascular diseases, diabetes, respiratory diseases and cancer.^3–6^ These diseases are common among homeless people. For example, a survey of homeless people in London and Birmingham found that the prevalence of cardiovascular disease was double that of housed people of the same age and sex, while prevalence of chronic obstructive pulmonary disease was ten times higher.^7^ The high prevalence of long-term respiratory disease and alcohol and drug problems may also make it difficult for people experiencing homelessness to recognise or act on COVID-19 symptoms.

In the UK, as in many countries, the government has provided advice and enacted new laws to protect the population from COVID-19. This includes advice to practice hand and respiratory hygiene, to only leave the house for very limited purposes, and to stay at home at all times if a household member has symptoms of COVID-19 (“self-isolation”). Many homeless people will be unable to follow these guidelines due to lack of handwashing facilities and lack of private accommodation. Some settings have shared bedrooms and bathrooms, and not all can be occupied during the day, whilst high turnover of residents may increase transmission risks.

### COVID-TRIAGE, COVID-PROTECT and COVID-CARE

In response to these concerns, some countries have developed plans to support homeless people during the COVID-19 pandemic. Publicly-funded programmes in New York City and Los Angeles County are using hotel rooms to shelter homeless vulnerable people, and establishing separate “medical sheltering” facilities for those with symptoms.^8,9^ Similarly, health and housing authorities in England have developed a plan to provide single room own-bathroom accommodation to homeless adults in England.^10^ This plan involves a triage process to identify people who either have symptoms of COVID-19, or are asymptomatic but at high risk of severe disease if they get COVID-19. People with symptoms will be offered ’COVID-CARE’ accommodation, where testing, isolation, observation, medical support and escalation to hospital are provided. People who are asymptomatic but at high risk will be offered ’COVID-PROTECT’ accommodation, which is designed to reduce the risk of becoming infected. Accommodation may be provided in hotels that are unoccupied during the pandemic or in single-room own-bathroom hostel accommodation. As of April 2020, the programme is at early stages of implementation across the UK. Asymptomatic people without specific vulnerabilities will receive information and advice. Figure 1 shows the envisaged intervention design. This study aims to estimate the potential impact of COVID-19 on the homeless population of England, and estimate the potential reduction in mortality and healthcare use that could be achieved by the intervention.

**Figure 1:**
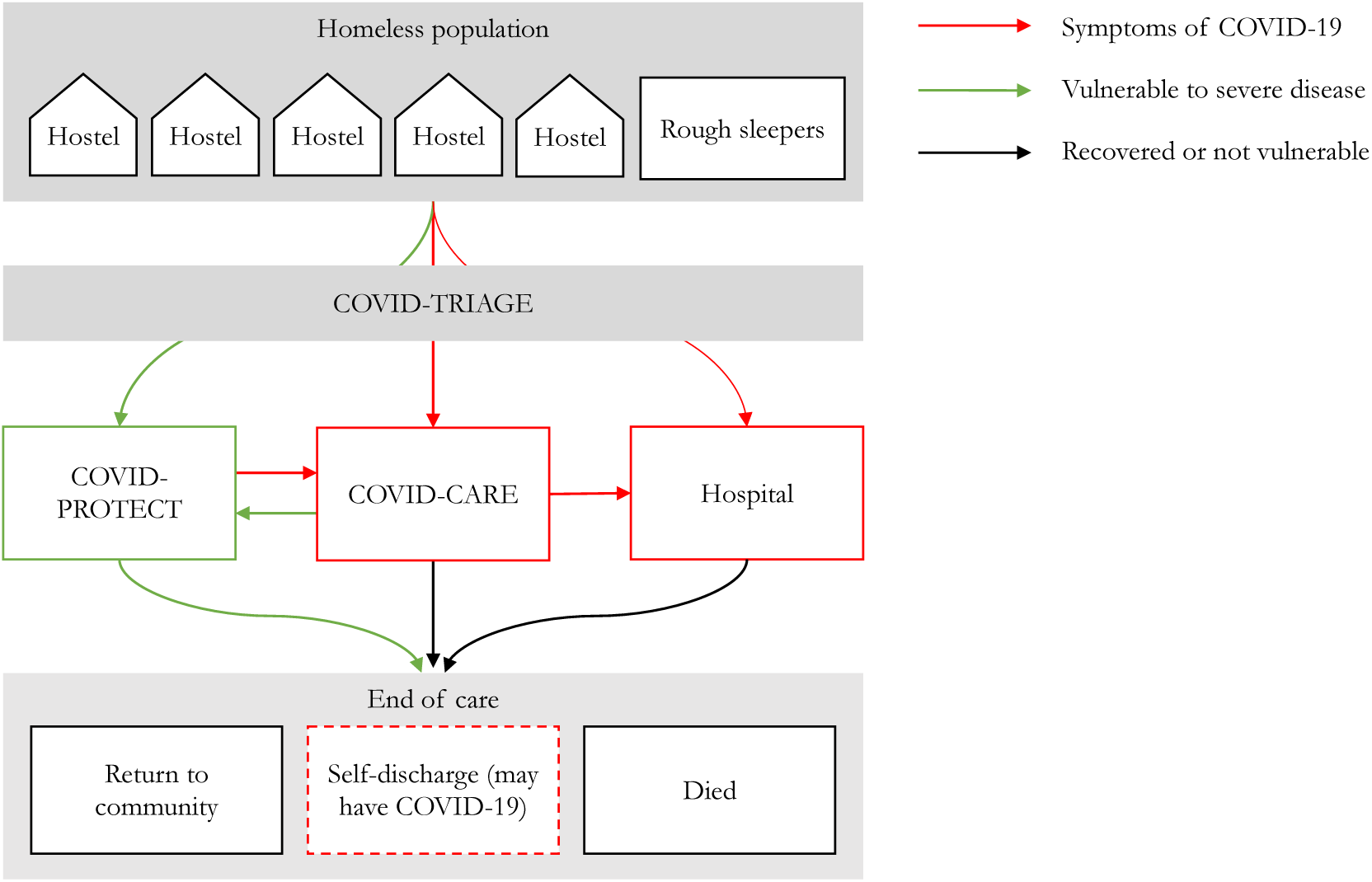
COVID-TRIAGE, COVID-PROTECT and COVID-CARE: diagram of an intervention to support homeless people during the COVID-19 pandemic

## Methods

We used a discrete-time Markov chain^11^ model to simulate stochastic daily transitions of individuals between four settings: the community (meaning living in a hostel or sleeping rough), COVID-PROTECT, COVID-CARE and hospital; and whether individuals were susceptible to COVID-19, exposed, infectious, or recovered (and immune, or died), following the SEIR (susceptible-exposed-infectious-recovered) approach^12^ to modelling of infectious diseases.

### Modelling COVID-19 among people experiencing homelessness

We assumed that transmission of COVID-19 among people experiencing homeless occurs within subgroups. For those living in hostels, we modelled transmission within hostels. A census of providers of accommodation for single homeless people suggested there were 35,817 people living in 1,065 hostels in England in 2019, and the median hostel size was 21 beds (IQR 12–38). There are limited estimates of the number of people sleeping rough, and we used an official government count^13^ in combination with an estimate of the number of people not captured by this count, producing an estimate of 10,748 people currently sleeping rough in England (see Supplementary Information for detail of this estimate). We did not have evidence of the structure of social mixing among people sleeping rough, and we created synthetic subgroups of sizes 1 to 100, where the group size was sampled with probabilities of the inverse of the size. This generated 530 subgroups of median size 8 (IQR 2–30), with approximately one-third of the population in 406 groups of size 1–33, one third in 80 groups of size 34–67, and the rest in 44 groups of size 67–100. We assumed homogeneous mixing within subgroups, but no mixing or transmission between them.

We assumed that people in both of these populations have a ’background’ risk of contracting COVID-19 from contact with staff or members of the general population. This background risk was based on the expected incidence in an existing model of COVID-19 transmission in the general population of the UK.^14^ The general population model forecasts a first wave of COVID-19 in the UK from March-June 2020, based on information and policies at the time of publication, with subsequent waves as policies are relaxed and reintroduced. New cases arising from this background incidence acted as seeds for transmission within hostels and the subgroups of people sleeping rough.

For each day of the model, we calculated a force of infection within each subgroup, based on the proportion of individuals that are infectious and values of R0. We assumed R0 for hostels to be 3.2 in the absence of any containment measures, based on a household secondary attack rate of 15% observed for COVID-19^15^ and the median size of hostels (21 beds), and 2.7 for people sleeping rough, based on the median value of R0 used in a model of COVID-19 transmission in the general population of the UK.^14^ We used published COVID-19 case series to create assumptions regarding the typical duration of the incubation and infectious periods (table 1), and assumed that these durations followed a gamma distribution across individuals.

**Table 1:** Numerical assumptions (base scenario)

| Assumption | Value | Source |
| --- | --- | --- |
| Number of people living in homeless hostels in England | 35,817 in 1,065 hostels | Number of beds in hostels for single homeless people, 2019<sup>16</sup> |
| Number of people sleeping rough in England | 10,748 | Official count of people sleeping rough (4,226),<sup>13</sup> plus an estimate of case ascertainment (40%) |
| Proportion of homeless people who are defined as vulnerable (aged 55+ or having a long-term condition) | 37% | Cross-sectional survey of 1,709 homeless people in London and Birmingham<sup>7</sup> |
| Incidence of COVID-19 in the general population of England | Varying by day and peaking in the first wave at 704 per million on 30 March 2020 | Most likely scenario in modelling of COVID-19 in the general population of the UK, accounting for non-pharmaceutical interventions such as staying at home<sup>14</sup> |
| Basic reproductive rate of COVID-19 in homeless hostels | 1.4 | Value of R0 required to achieve cumulative incidence of 8% at the end of April 2020 |
| Basic reproductive rate of COVID-19 among people sleeping rough | 1.4 | As above |
| Basic reproductive rate of COVID-19 in COVID-CARE and COVID-PROTECT | 0.75 | Assuming strong infection control measures |
| Probability of being identified (if eligible for COVID-CARE or COVID-PROTECT) by COVID-TRIAGE workers | 70% | Estimate based on early programme experience in London |
| Probability of accepting COVID-CARE or COVID-PROTECT when offered | 80% | Estimate based on early programme experience in London |
| Risk of self-discharge from COVID-CARE or COVID-PROTECT over two weeks | 8% | Based on the risk of self-discharge during hospital admissions (which was 8% in a study of 3,222 homeless inpatients in England<sup>17</sup>), and early programme experience in London |
| Infection fatality ratio for patients aged under 55 and without a long-term condition | 0.6% | Based on estimates of the IFR from existing observational studies in the general population, after adjustment for biases relating to under-ascertainment and delays from onset to death. See Supplementary Information for detail. |
| Infection fatality ratio for patients aged 55+ or who have a long-term condition | 4.8% | Based on an estimated relative risk of death associated with older age or underlying health conditions of 8.2. See Supplementary Information for detail. |
| Proportion of COVID-19 in each illness severity group | See table 3 | Proportions are guided by published COVID-19 case series and our estimates of the overall infection fatality ratio. See table 3 and Supplementary Information for detail. |
| Fatality ratio by illness severity group | See table 3 | Assuming that most deaths occur in the 'critical' group, based on data from China<sup>18</sup> and the UK,<sup>19</sup> with some adjustment for due to delays to death beyond the end-point of these analyses |
| Mean durations in the exposed and infectious compartments | Latent (pre-infectious): 2.5 days<br>Pre-symptomatic infectious: 2.5 days<br>Symptomatic and infectious: 4 days | Estimates based on a modelling study using data from 77 transmission pairs<sup>20,21</sup> |
| Mean duration between onset of symptoms and admission to hospital or ITU | 7 days | Median time from onset of symptoms to hospitalisation in 41 patients in China,<sup>22</sup> and median time from onset of symptoms to hospitalisation for 138 patients in China<sup>23</sup> (both 7 days, IQR 4-8) |
| Mean duration of hospital or ITU admission | 11 days | Median length of hospital stay for 191 patients in China (11 days, IQR 7-14)<sup>6</sup> |
| Mean duration between acquisition of COVID-19 and end of disease (recovery or death) | 23 days | Median time from onset of symptoms to death for 54 patients who died in China (18.5 days, IQR 15-22) plus estimate of pre-symptomatic period<sup>6,24</sup> |
| Incidence of influenza like illness due to causes other than COVID-19, assumed to apply to individuals in all settings | 6.3 per 100 person-weeks | Cohort study of the general population in the UK, 2012-2013<sup>25</sup> |

Reports from hostels in London suggest that hostel staff and residents are taking measures to reduce social contact and few residents have become unwell. Testing for COVID-19 among homeless people in London in April 2020 found that 8% of those tested were positive. This information led us to revise our estimates of R0 down to 1.4, as this value produced a cumulative incidence of approximately 8% at the end of April.

We classified cases as asymptomatic, mild (requiring basic clinical observation), moderate (requiring some care, such as oxygen), severe (requiring care in hospital), or critical (requiring treatment in an ITU), with higher mortality risks for more severe cases. Our assumed probabilities of each severity shown in table 3. We divided participants into those with specific vulnerability to severe disease (based on age or comorbidity) and those who did not. The ’vulnerable’ group had a higher risk of more severe disease. Individuals with different disease severities, including asymptomatic cases, were assumed to have equal infectiousness.

We ran the model for 28 weeks, from 29 January to 12 August 2020, with the intervention first available on 1 March.

### Modeling the demand and impact of COVID-TRIAGE, COVID-PROTECT, and COVID-CARE

In our model, individuals who are asymptomatic (i.e. susceptible to COVID-19 or pre-symptomatic) and vulnerable to severe disease are offered COVID-PROTECT steadily over a period of 28 days. We assumed that 70% of eligible individuals are identified, of which 80% accept the intervention when offered (with the rest remaining in hostels or sleeping rough). Individuals who develop symptoms, either due to COVID-19 or other respiratory illnesses, are offered COVID-19 on the day they become symptomatic. To allow for this, we included typical durations of ’pre-symptomatic’ and ’symptomatic’ infectiousness in our model, in addition to the standard SEIR statuses. Susceptible individuals in COVID-PROTECT or COVID-CARE may acquire COVID-19 either through mixing with staff (with the same ’background’ risk level as hostel and rough sleeping populations defined by the general population incidence, as described above) or a force of infection specific to COVID-PROTECT or COVID-CARE. Those who developed symptoms due to causes other than COVID-19 are returned to their origin after two days, on receiving test results. We assumed that moderate and severe cases in the community (i.e. those not identified by COVID-TRIAGE or who declined COVID-CARE) are admitted to hospital, while critical cases are admitted to an ITU. In COVID-CARE, mild and moderate cases are treated without hospital admission, severe cases are transferred to hospital, and critical cases to an ITU. If people develop symptoms while in COVID-PROTECT (which may be due to COVID-19 or other respiratory diseases), they are transferred to COVID-CARE. Our numerical assumptions are summarised in Table 1, with further detail in Supplementary Information.

### Scenario analysis

We reported key results for eight scenarios in which the intervention model varied. The main two scenarios were ’do nothing’ and the ’base intervention’ scenario, which include the assumptions we considered most likely at the time of publication. We also ran the model in six other scenarios, representing different implementation and policy choices. These scenarios are listed in table 2.

**Table 2:** Parameters for scenarios

| Scenario |  | Description | R0: Hostels<br>(up to 3<br>June) | R0: Sleeping<br>rough (up<br>to 3 June) | R0: Sostels<br>(after 3<br>June) | R0: Sleeping<br>rough (after<br>3 June) | Proportion<br>accepting | Testing<br>available in<br>COVID-CARE | COVID-PROTECT<br>eligibility | Limit on<br>COVID-PROTECT |
| --- | --- | --- | --- | --- | --- | --- | --- | --- | --- | --- |
| 1 | Base (do<br>nothing) | Best estimate in<br>the absence of<br>an intervention | 1.4 | 1.4 | 1.4 | 1.4 | 0% | - | - | - |
| 2 | Base (inter-<br>vention) | Best estimate<br>with COVID-PROTECT and<br>COVID-CARE | 1.4 | 1.4 | 1.4 | 1.4 | 80% | Yes | Age 55+<br>and/or LTC | None |
| 3 | No testing | Scenario 2, but<br>people other<br>respiratory<br>diseases are not<br>discharged on<br>negative results | 1.4 | 1.4 | 1.4 | 1.4 | 80% | No | Age 55+<br>and/or LTC | None |
| 4 | No triage | Scenario 2, but<br>everyone is<br>offered COVID-PROTECT and<br>beds are limited<br>to 9,000 | 1.4 | 1.4 | 1.4 | 1.4 | 80% | Yes | Everyone | 9,000 beds |
| 5 | Lift<br>restrictions<br>on 3 June<br>(do<br>nothing) | Scenario 1, but<br>with<br>containment<br>measures lifted<br>on 3 June | 1.4 | 1.4 | 3.2 | 2.7 | 0% | - | - | - |
| 6 | Lift<br>restrictions<br>on 3 June<br>(interven-<br>tion) | Scenario 2, but<br>with<br>containment<br>measures lifted<br>on 3 June | 1.4 | 1.4 | 3.2 | 2.7 | 80% | Yes | Age 55+<br>and/or LTC | None |
| 7 | No<br>restrictions<br>(do<br>nothing) | Scenario 1, but<br>with no<br>containment<br>measures | 3.2 | 2.7 | 3.2 | 2.7 | 0% | - | - | - |
| 8 | No<br>restrictions<br>(interven-<br>tion) | Scenario 2, but<br>with no<br>containment<br>measures | 3.2 | 2.7 | 3.2 | 2.7 | 80% | Yes | Age 55+<br>and/or LTC | None |

**Table 3:** Assumptions relating to case severity and infection fatality ratios

|  | Proportion of cases |  |  | IFR per 10,000 |  | Healthcare use |  |
| --- | --- | --- | --- | --- | --- | --- | --- |
|  | Non-vulnerable | Vulnerable | Overall | Non-vulnerable | Vulnerable | Community | COVID-CARE |
| Asymptomatic | 40% | 18% | 29% | 0 | 0 | None | None |
| Mild | 45% | 30% | 42% | 1 | 1 | None | CARE |
| Moderate | 10% | 25% | 16% | 25 | 25 | Hospital | CARE |
| Severe | 4% | 20% | 9% | 200 | 200 | Hospital | Hospital |
| Critical | 1% | 7% | 4% | 6,000 | 6,000 | ITU | ITU |
| Total | 100% | 100% | 100% | 59 | 484 |  |  |

IFR = infection fatality ratio

### Sensitivity analysis

We conducted sensitivity analyses for five variables representing the quality of the intervention: (1) the value of R0 in COVID-PROTECT; (2) the value of R0 in COVID-CARE; (3) the proportion of homeless people who are identified for triage; (4) the proportion who accept when offered COVID-PROTECT or COVID-CARE; (5) the proportion that self-discharge. We also conducted sensitivity analysis for two disease-related factors: (6) infection mortality rates, and (7) where R0 is low (1.1 rather than 1.4 for both people living in hostels and people sleeping rough) and high (3.2 for people living in hostels and 2.7 for people sleeping rough).

For each point estimate, we ran the model 1,000 times and reported the 0.025, 0.5 and 0.975 quantiles. Where we plotted time series, we selected the ’median run’ as the model producing the median number of cumulative deaths.

We also tested some longer-term scenarios where transmission is lower in the short term, with results in Supplementary Information.

We used R version 3.6.2^26^ to build the model, and have published the code (https://github.com/maxeyre/Homeless-COVID-19). We have also created an online interface (https://care-protect.shinyapps.io/covid-19/) that allows users to adjust model inputs and assumptions and view results (in development at the time of pre-print publication).

## Results

### Cases of COVID-19

In the ’do nothing’ scenario, the model suggests that 15,448 cases of COVID-19 could occur among people living in hostels and sleeping rough combined, with cumulative incidence of 34%. Among the 1,595 clusters (1,065 hostels and 530 clusters of people sleeping rough), 45% are unaffected (mean size 12 individuals), 33% have some cases but fewer than half of residents contracting COVID-19 (mean size 39 individuals), and 22% have more than half contracting COVID-19 (mean size 51 individuals).

In the base scenario where COVID-PROTECT and COVID-CARE are implemented, only 8% of clusters have more than half of individuals contracting COVID-19. This is because outbreaks with high levels of exposure for all individuals are avoided by removing symptomatic cases and placing them into COVID-CARE. In the base intervention scenario, 18% of individuals contract COVID-19 in the community, and a further 3% in COVID-PROTECT or COVID-CARE. This is a cumulative incidence of 21%, or 9,934 cases, representing 5,514 avoided cases when compared to the ‘do nothing’ scenario.

### Mortality due to COVID-19

Given our assumptions of the mortality risk associated with COVID-19 and the proportion of the population that is vulnerable to severe disease, we estimated 364 deaths in the ’do nothing’ scenario and an infection fatality ratio of 2.4%. The model suggested that the intervention could reduce the number of deaths to 200, representing a potential to avoid 164 deaths. The infection fatality ratio is 2.0%, which is lower because COVID-PROTECT reduces risk of COVID-19 among vulnerable people more than amongst other people.

**Figure 2:**
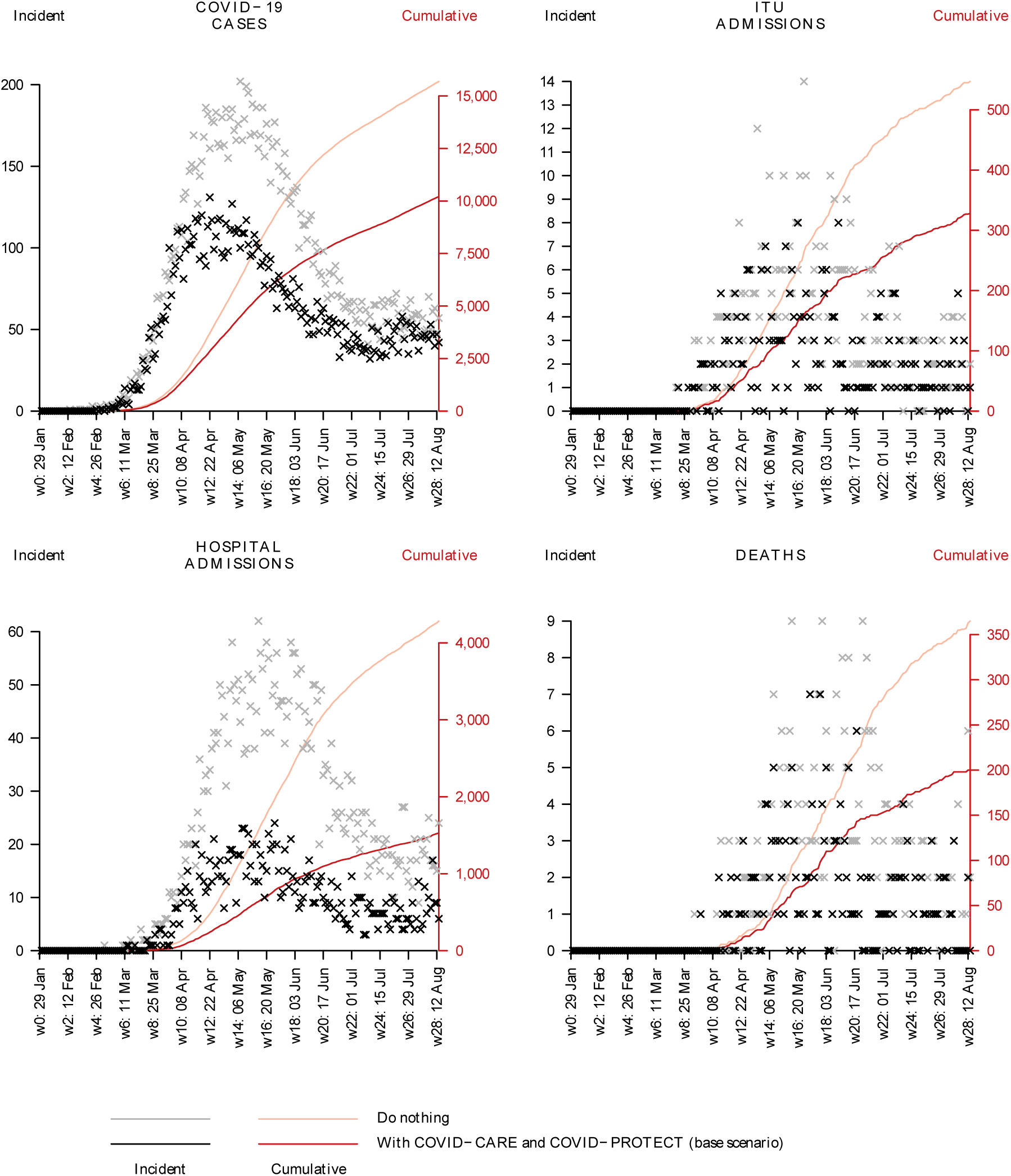
COVID-19 cases, deaths, and hospital admissions (charts show a single run that resulted in the median number of deaths)

### Demand for COVID-PROTECT and COVID-CARE

Demand for COVID-CARE follows the incidence of COVID-19 in the community, peaking at 1,366 beds on 5 May. Demand then reduces as residents recover, return to the community or COVID-PROTECT, or self-discharge. In our base intervention scenario, there is low incidence of COVID-19 and most individuals in COVID-CARE have symptoms due to other causes. Demand for COVID-PROTECT increases over the period of recruitment (assumed to be 28 days) to 9,290 beds, then oscillates as people self-discharge, become symptomatic and are transferred to COVID-CARE, or are transferred back from COVID-CARE or hospital on recovery.

**Figure 3:**
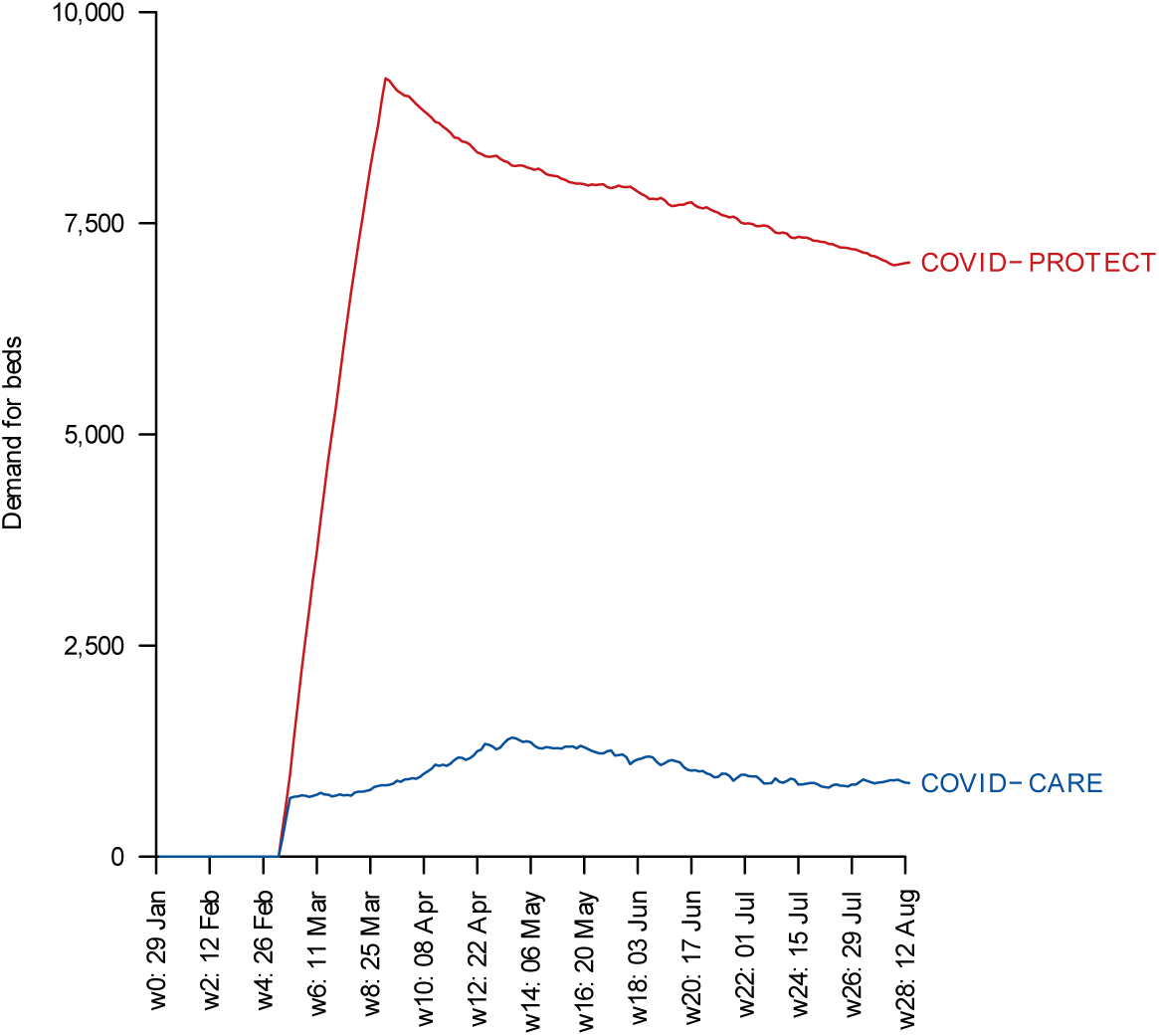
Modelled demand for beds in COVID-PROTECT and COVID-CARE (base case scenario)

### Hospital use

In the ’do nothing’ scenario, homeless people in England are admitted to hospital for non-ITU treatment 4,074 times and a further 572 are admitted to an ITU. Individuals in our model are only admitted to hospital once. In the ’base scenario’, hospital admissions reduced to 1,450 and ITU admissions reduced to 324. This represents a potential to avoid 2,624 hospital admissions and 248 ITU admissions.

### Scenario analysis

The ’no testing’ scenario, in which there is no access to testing within COVID-CARE facilities, substantially increases peak demand for COVID-CARE beds. This is because, without testing, all symptomatic individuals may require admission for two weeks, whether they have COVID-19 or a common respiratory infection. There are also increases in the number of COVID-19 cases and deaths because susceptible individuals are admitted to COVID-CARE where there may be greater risk of infection than in the community. The ’no triage and limited capacity’ scenario (in which all asymptomatic individuals are eligible for COVID-PROTECT but capacity is limited) results in a greater number of deaths, hospital and ITU admissions than the base case scenario, and with similar demand for COVID-PROTECT and COVID-CARE. Summaries from the scenario analysis are shown in Table 3. In scenarios where ‘normal mixing’ resumes in the homeless population after 3 June 2020, a second peak of COVID-19 occurs (see Supplementary Information for time series of the number of cases under each scenario).

**Table 3:** Analysis of intervention design scenarios. Median (2.5%, 97.5% quantile) from 1,000 model runs

| Scenario | Peak PROTECT demand | Peak CARE demand | Total COVID-19 cases | Number of deaths | Number of hospital admissions | Number of ITU admissions |
| --- | --- | --- | --- | --- | --- | --- |
| Base (do nothing) | - | - | 15,448<br>(14,795-16,001) | 364<br>(332-420) | 4,074<br>(3,847-4,279) | 572<br>(526-640) |
| Base (intervention) | 9,290<br>(9,133-9,421) | 1,366<br>(1,308-1,434) | 9,934<br>(9,556-10,546) | 200<br>(171-222) | 1,450<br>(1,384-1,557) | 324<br>(284-349) |
| No testing (intervention) | 7,998<br>(7,778-8,124) | 4,327<br>(4,247-4,408) | 11,436<br>(10,578-11,960) | 268<br>(231-302) | 1,691<br>(1,524-1,832) | 430<br>(388-475) |
| No triage (intervention) | 9,916<br>(9,845-9,993) | 1,406<br>(1,345-1,470) | 9,820<br>(9,249-10,520) | 223<br>(186-251) | 1,579<br>(1,461-1,705) | 360<br>(321-396) |
| Lift restrictions on 3 June (do nothing) | - | - | 22,378<br>(21,422-23,142) | 462<br>(416-509) | 5,528<br>(5,185-5,778) | 782<br>(725-846) |
| Lift restrictions on 3 June (intervention) | 9,290<br>(9,133-9,421) | 1,593<br>(1,481-1,758) | 15,696<br>(14,988-16,589) | 246<br>(224-275) | 2,068<br>(1,943-2,231) | 438<br>(395-486) |
| No restrictions (do nothing) | - | - | 33,340<br>(32,364-34,116) | 828<br>(767-879) | 8,955<br>(8,581-9,195) | 1,274<br>(1,203-1,333) |
| No restrictions (intervention) | 9,257<br>(9,088-9,439) | 2,914<br>(2,738-3,062) | 25,661<br>(24,783-26,467) | 490<br>(454-534) | 3,854<br>(3,693-4,019) | 761<br>(704-817) |

### Sensitivity analysis

Sensitivity analyses highlighted the importance of transmission in COVID-PROTECT. Increases in the value of R0 in this setting led to large increases in the number of deaths and hospital admissions. R0 in COVID-PROTECT has a large impact because the residents are vulnerable to severe disease. The proportion of eligible individuals that are identified or accept the interventions has a direct relationship with the benefits of the interventions. Improvements in acceptability (reflected in greater proportions accepting the interventions or fewer self-discharging) have a greater impact on deaths and healthcare use than on the number of cases (which are not greatly affected in any of the sensitivity analyses), reflecting targeting of COVID-PROTECT towards more vulnerable people.

**Figure 4:**
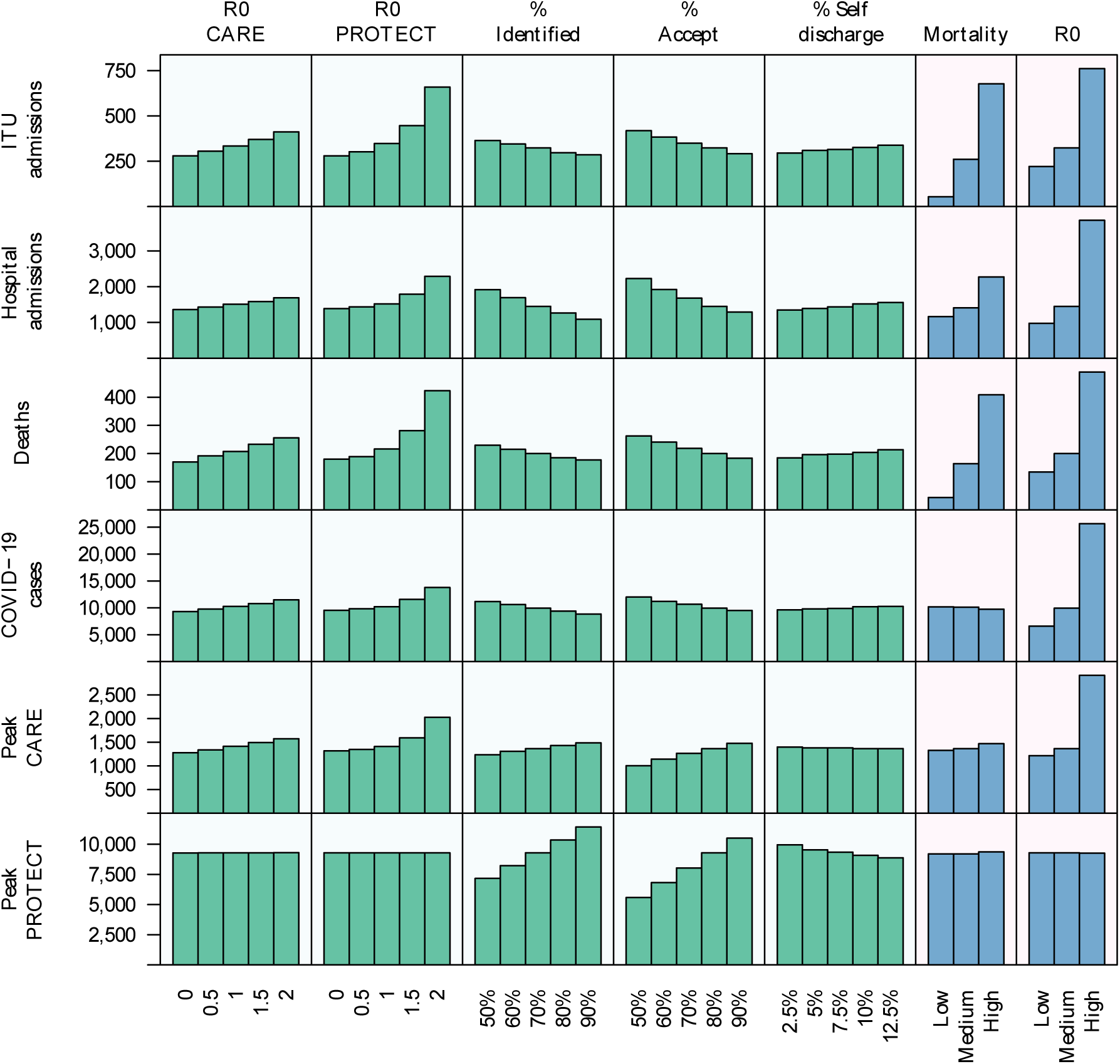
Results of sensitivity analysis (showing median values from 1,000 model runs)

## Discussion

### Key findings

Over the first wave of the COVID-19 epidemic, COVID-PROTECT and COVID-CARE could reduce deaths among homeless people in England by 45%, hospital admissions by 64% and ITU admissions by 43%, compared with a ’do nothing’ scenario. Our results highlight the importance of measures to reduce transmission within COVID-PROTECT and COVID-CARE, the benefits of triage on the basis of vulnerability (if capacity is limited), and the reductions in peak COVID-CARE demand and deaths that could be reduced through PCR testing.

### Strengths and limitations

To our knowledge this model is the first estimate of potential COVID-19 transmission in a homeless population. The impact of COVID-19 on people experiencing homelessness in the United States has been estimated, though this model uses a fixed ’peak infection’ of 40% and does not model transmission.^9^ We used a dynamic transmission model that allowed us to estimate the benefits of isolating vulnerable and infectious individuals. We used real-time data from the homeless population of London to calibrate our assumptions about transmission. Our scenario analysis identified key success factors for the intervention. We sought to account for real-world complexities including behavioural factors (such as non-acceptance of COVID-CARE or COVID-PROTECT and self-discharge), asymptomatic and pre-symptomatic cases being admitted to COVID-PROTECT and thereby increasing transmission in this setting, respiratory symptoms due to causes other than COVID-19 (which may lead to people who are susceptible to COVID-19 being admitted to COVID-CARE), and limits to accommodation and COVID-19 testing capacity.

The main limitations of the model relate to uncertainties regarding COVID-19’s epidemiological and clinical characteristics, operational issues such as bed capacity, and residents’ behaviour within COVID-CARE and COVID-PROTECT. We used published evidence to support our assumptions where possible, and sought feedback from healthcare workers and programme managers who are starting to implement COVID-CARE and COVID-PROTECT in some regions. For example, we adjusted values of R0 to try and make the model results reflect early data, and we estimated the proportion accepting either type of accommodation when offered, and the proportion self-discharging, and tested these in sensitivity analysis. However, true values may differ and are also influenced by the design and management of the intervention.

We were unable to account for limits to the availability of hospital and ITU beds, and to what extent this may affect mortality rates of cases remaining in COVID-CARE or in the community. Capacity in acute hospitals may vary by region as well as over time, particularly as temporary hospitals are established in large urban areas,^27^ but this is not reflected within our model. Nor did we account for patients who might be too frail to be appropriate candidates for invasive ventilation, and for whom palliative care may be more appropriate.

There is uncertainty in the infection fatality ratio of COVID-19, as well as effects of age, comorbidities, and other patient factors. As shown in our sensitivity analysis, the infection fatality ratio has a large bearing on the expected number of deaths. One theory is that the risk of death after being infected by COVID-19 risk approximates to an individual’s annual mortality risk from all other causes.^28^ Cohort studies of people experiencing homelessness have found standardised mortality ratios of three to six,^29–32^ which may suggest that the infection fatality ratio for this population would be three to six times the general population. Our model gave an infection fatality ratio of 2.4%, which is approximately four times an estimate of 0.6% for the general population,^33^ which supports our assumptions for COVID-19 mortality risk among homeless people.

We did not stratify our input parameters by sex, although around 80% of single homeless people are male.^7,34^ There is some evidence that male sex might be associated with increased mortality from COVID-19 (for example in the data from China, the naive case fatality rate was estimated at 2.7% in males and 1.8% in females).^18^ Tobacco smoking may also increase mortality risk beyond what might be expected based on co-morbidities alone,^35^ which is concerning given the high prevalence of smoking among homeless people.

We focused on a period that is expected to represent the ’first wave’ of COVID-19. However, the disease may be transmitted over a longer period, particularly with intermittent suppression or mitigation measures.^1^ Deaths from COVID-19 averted over this period may be delayed if the intervention is ended in summer 2020, and if individuals are not discharged to suitable longer-term accommodation with individual rooms and bathroom facilities. Linked to this, our model does not allow for waning immunity; which appears reasonable over three months^36–38^ but immunity over longer periods is unknown.

### Implications and recommendations for practice

Our sensitivity analysis shows that the proportion who self-discharge and the proportion that accept COVID-PROTECT or COVID-CARE affect overall mortality. Early programme experience suggests limited numbers self-discharge from these facilities, but this may vary. The acceptability of the intervention will affect both the likelihood that eligible individuals will accept the intervention and the risk of self-discharge. Measures to improve acceptability may include (a) provision of clear information prior to admission, (b) avoidance of a ’detox’ or ’abstinence’ approach to alcohol, drugs and smoking, with involvement of drug and alcohol services, and access to substitution therapies and other harm reduction services, (c) involvement of mental health services and support, (d) provision of televisions, smartphones or tablets to improve residents’ access to entertainment, social connection, support from staff and peers, and to facilitate clinical observations, (e) involvement of experienced staff including trained peer support workers who can help COVID-PROTECT and COVID-CARE meet residents’ needs, (f) assurances that placement in COVID-PROTECT or COVID-CARE will not lead to people losing their current accommodation.

The comparison between the ’do nothing’ scenario and the base intervention scenario suggests that outbreaks within hostels or social groups can be avoided by moving people with symptoms into accommodation where isolation is possible. If this is not done, the secondary attack rate within these settings can be very high, with more than 90% of residents affected in many hostels. With low incidence in the general population, larger hostels are likely to be at higher risk of an outbreak due to more contact with staff and a high probability of one case. Using our assumptions, the benefit of reducing outbreaks in the community outweighs the risks of transmission within COVID-PROTECT and COVID-CARE. This is reflected in data from the United States, where COVID-19 testing in homeless shelters in the United States found that shelters where two or more COVID-19 cases were reported in the preceding two weeks had much higher prevalence than shelters with one or zero reported prior cases (37% compared to 5%),^39^ suggesting that outbreaks in these settings are an important element of COVID-19 risk for homeless populations.

Our results also show the importance of minimising transmission of COVID-19 within COVID-PROTECT. This includes avoidance of close contact for both residents and staff, promotion of respiratory and hand hygiene, and personal protective equipment where recommended. Methods of symptom-monitoring, including staff carrying out daily check-ins and mechanisms to help residents self-report a new cough or fever, may help staff to isolate residents with possible COVID-19 and to identify outbreaks. Managing an outbreak in COVID-PROTECT is likely to entail practical challenges. Nevertheless, identification and control of outbreaks is likely to be more feasible in this setting compared to standard hostel accommodation with shared facilities.

If programme capacity is limited, the use of a triage process to prioritise those most at-risk from COVID-19 for COVID-PROTECT accommodation is likely to reduce mortality, by targeting resources towards those most likely to benefit. Effective triage is also important to ensure that only asymptomatic individuals are admitted to COVID-PROTECT, which is likely to reduce risks of transmission to other residents.

## Conclusions

Compared to the general population, homeless people are at higher risk from COVID-19 due to increased exposure in the community and greater vulnerability to severe disease. Our modelling shows that deaths and hospital use can be reduced through separation of infected and vulnerable people in COVID-CARE and COVID-PROTECT. There remains considerable uncertainty in the intervention structure and impact and it will need ongoing monitoring and evaluation.

## Data Availability

All data and code has been made publicly available.

https://github.com/maxeyre/Homeless-COVID-19

## Acknowledgments

Homeless Link provided data on the number of hostels in England and the number of beds in each hostel. Nicholas Davies at the London School of Hygiene and Tropical Medicine provided detailed results from a model of COVID-19 transmission in the general population of the UK.

